# Questioning the association of the *STMN2* dinucleotide repeat with ALS

**DOI:** 10.1101/2022.04.30.22273566

**Authors:** Jay P. Ross, Fulya Akcimen, Calwing Liao, Dan Spiegelman, Ben Weisburd, Nicolas Dupré, Patrick A. Dion, Guy A. Rouleau, Sali M. K. Farhan

## Abstract

Lowered expression of *STMN2* is associated with TDP-43 pathology in amyotrophic lateral sclerosis (ALS). Recently, the number of dinucleotide CA repeats in an intron of the *STMN2* gene was reported to be associated with increased risk for ALS. Here, we used a case-control cohort of whole genome sequencing (WGS) as well as WGS from populations in the gnomAD cohort to attempt to replicate this proposed association. We find that repeats well above the previously reported pathogenic threshold of 19 are commonly observed in unaffected individuals across different populations. Further, we did not observe an association between longer *STMN2* CA repeats and ALS phenotype. In summary, our results do not support a role of *STMN2* CA repeats towards ALS risk.

**Disclosures:** None to disclose

## Introduction

Altered Stathmin-2 (*STMN2*) gene expression and isoform usage have recently been implicated in amyotrophic lateral sclerosis (ALS) cellular pathology^1,2^. Upon decrease in *TARDBP* gene expression or TDP-43 protein function, the *STMN2* transcript becomes truncated and produces a non-functional stathmin-2 protein. This dysfunction results in altered neural response to cell damage and reduced ability to regrow axons.

Recently, an intronic dinucleotide CA repeat between exons 3 and 4 of the *STMN2* gene was reported to be associated with risk for ALS^3^. Specifically, alleles longer than 19 CA repeats were observed to impart an increased risk for ALS, and those carrying a 24-repeat allele alongside another long allele had the highest risk for ALS.

In the current study, we tested the *STMN2* CA repeat in a case-control cohort of European genetic background, as well as in genomes from various populations in gnomAD^4^. We observed carriers of *STMN2* CA repeats well beyond the previously reported pathogenic repeat threshold in both case-control and gnomAD cohorts, and we did not reproduce the association between expanded *STMN2* repeats and ALS. While *STMN2* may be an important aspect of ALS cellular pathology, the dinucleotide repeat in *STMN2* does not appear to impart significant risk to develop ALS.

## Methods

### Sample cohorts and genome sequencing

ALS patients and unaffected controls were recruited in clinics across Québec, Canada. 154 ALS patients (average age of ALS onset 59.7 ± 11.7 years, male:female ratio 1.68) were included. 216 unaffected controls (average age 67.8 ± 13.3 years, male:female ratio 0.56), originating from separate studies on essential tremor or restless legs syndrome were included. While the latter cohort may have been affected for their respective phenotypes, no clinical description of motor neuron degeneration symptoms or neurodegenerative dementias was noted. The study populations in gnomAD were used as an external dataset^4^. All individuals included in the study gave written informed consent in accordance with the appropriate ethics review boards.

Whole genome sequencing (WGS) was performed on Illumina HiSeq X-Ten and Illumina Novaseq6000 sequencers at the Génome Québec Centre d’Expertise et de Services. Bioinformatic analyses were performed on the Béluga cluster of Compute Canada and Calcul Québec, using DRAGEN Bio-IT v3.8 (Illumina, Inc). After alignment to the hg38 human reference genome, an average depth of 34.1X. Whole genome sequencing for gnomAD populations was performed as previously described^4^.

### Estimation of STMN2 CA repeat length from WGS data

Expansionhunter v4.0.2^5^ was used to calculate the number of CA repeats in the locus previously reported^3^. Options used in ExpansionHunter were as follows: ReferenceRegion: chr8:79641628-79641672, VariantType: “Repeat”, LocusStructure: “(CA)*”. The reported sex of an individual was used as an input option; sex was imputed from WGS data if reported sex was not available.

### Statistical analyses

All statistical tests were performed using R v4.0.3. Fisher’s exact test (fisher.test) was used to test for differences in repeat length between cases and controls and to obtain odds ratios. A Cochran-Mantel-Haenszel (CMH) test (mantelhaen.test) was used to incorporate the current results with those of the previous Theunissen study^3^. All reported p-values are uncorrected. The “tidyverse” R package was used to generate plots and format data^6^. Per-sample allele lengths are reported in Supplemental Table S2.

## Results

The *STMN2* CA repeat was successfully genotyped by Expansionhunter in 153 ALS and 207 controls. No allele combination with the current case-control cohort suggested an association of long or long-with-24-repeats (L/L with 24CA) with the ALS phenotype (Table 1). While there was a nominally significant p-value of L/L with 24CA using the CMH test combining allele counts from the current cohort and the Australian cohort from the previous study (p = 0.041), this result does not pass the multiple-testing correction threshold (α = 0.05/10; p = 0.005). The longest repeats were more often observed in female samples, and the largest repeats were observed in female control samples (Table S2).

**Table 1.** Replication results of Theunissen et al associations of *STMN2* CA repeat lengths and ALS phenotype.

| Genotype | Status |  | Fisher's Exact Test |  |  | CMH test |  |  |
| --- | --- | --- | --- | --- | --- | --- | --- | --- |
|  | ALS | Controls | p-value | OR | 95_CI | p-value | OR | 95_CI |
| Long/Long (L/L) | 84 (54.9%) | 123 (59.4%) | 0.4504 | 1.20 | 0.77-1.87 | 0.1775 | 1.19 | 0.93-1.53 |
| L/L (with 24 CA) | 59 (38.6%) | 77 (37.2%) | 0.8263 | 0.94 | 0.60-1.48 | 0.0407 | 1.44 | 1.02-2.03 |
| L/L (without 24 CA) | 25 (6.94%) | 46 (12.8%) | 0.1819 | 1.46 | 0.82-2.62 | 0.9179 | 0.97 | 0.75-1.27 |
| Long/Short (L/S) | 44 (12.2%) | 62 (17.2%) | 0.8163 | 1.06 | 0.65-1.72 | 0.0935 | 0.79 | 0.61-1.03 |
| Short/Short (S/S) | 25 (6.94%) | 22 (6.11%) | 0.1167 | 0.61 | 0.31-1.18 | 0.2504 | 1.36 | 0.84-2.20 |

Tests of association were performed using either the Fisher's Exact Test or the Cochran-Mantel-Haenszel (CMH) test. P-values are reported uncorrected. Counts and percentages of individual carriers are listed for ALS and Control cohorts for each combination of allele length. Odds ratio (OR) and 95% confidence intervals (95\_CI) are given separately for each test and combination.

Notably, *STMN2* CA repeats much longer than the previously reported ALS-associated threshold were frequently observed in the gnomAD genomes populations (Figure S1). Repeat lengths as long as 89 were observed in the Non-Finnish European cohort, which is likely the closest match to both the current case-control cohort and the previously reported cohort^3^. The frequency of the different allele combinations as in Figure 1 varied slightly between gnomAD populations (Table S1).

**Figure 1.**
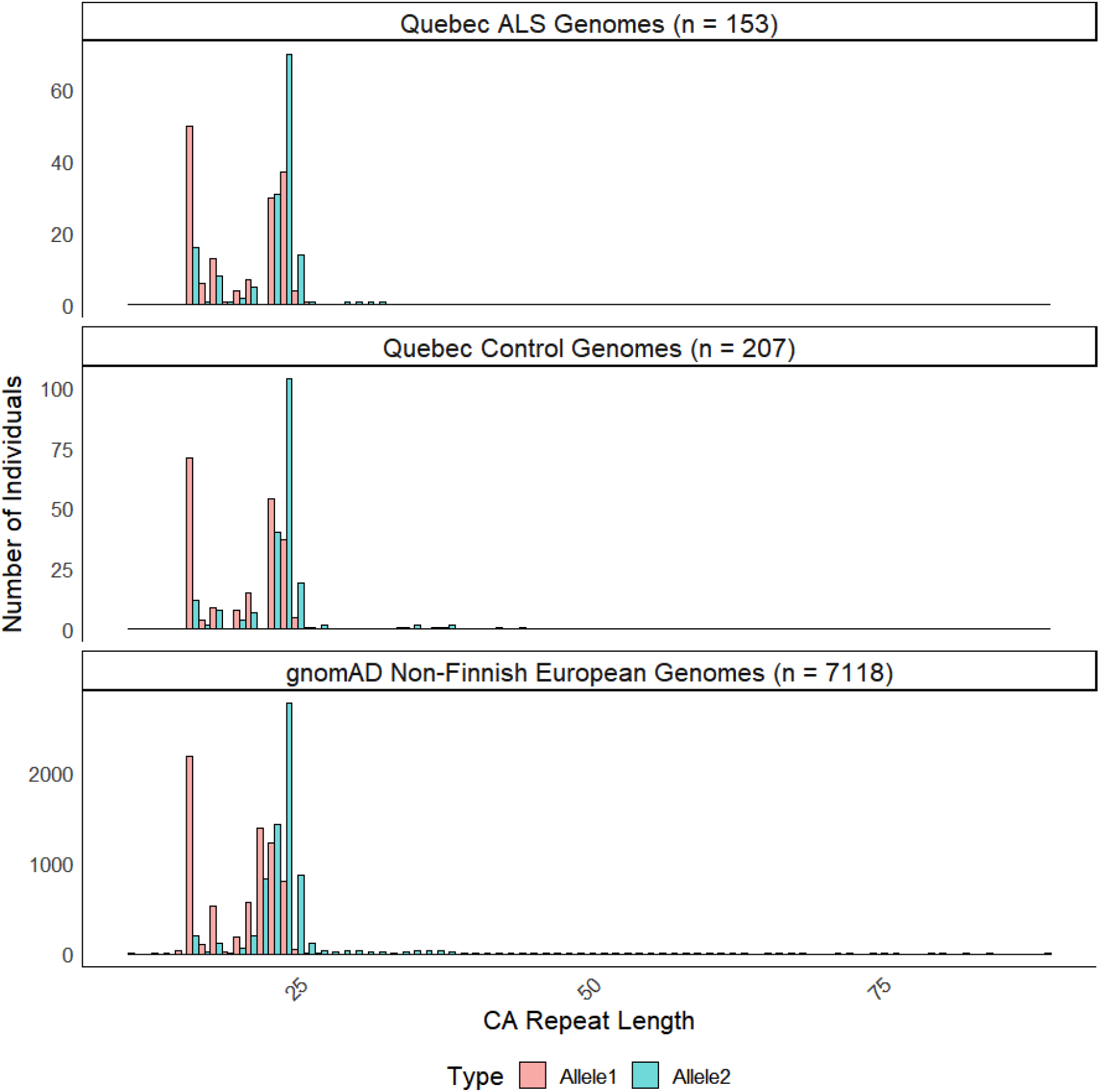
Distributions of *STMN2* CA repeat length calculated from ALS, control, and Non-Finnish European (NFE) gnomAD genomes. Expansionhunter v4.0.2 software was used to call STMN2 CA repeat lengths from genomes aligned to the hg38 human reference genome. Sample sizes for each respective group are listed above each panel. Alleles are noted as the shorter and longer allele per individual (Allele1 and Allele2) in red and blue, respectively.

## Discussion

Our study employed WGS data to accurately estimate the *STMN2* CA repeat length, and we observed very large repeats above the purportedly pathogenic threshold in phenotypically normal individuals. We did not observe an association between longer alleles and risk for ALS, nor did we replicate the necessity of the 24-repeat allele for this association.

The previous study reported a trend of large *STMN2* CA repeat length with decreased expression of *STMN2*^3^. However, this trend was not statistically significant. Further, it is unclear whether larger repeats are linearly associated with decreased *STMN2* levels, or whether the decrease is comparable to that resulting from *TARDBP* variation or lowered *TARDBP* expression^1,2^. While the expression level and pathological truncation of *STMN2* are important in ALS and TDP-43 pathology, our current results refute the association of the *STMN2* CA dinucleotide repeat with the disease. We recommend the use of unbiased genome-wide screens for modifiers of *STMN2* expression level in future studies to potentially identify new variants and genes relevant to ALS.

Our study had certain limitations. The case-control cohort was ascertained based on criteria external to the current study. Namely, ALS cases were selected based on sporadic family history and the absence of strongly penetrant ALS-associated variants. Controls were not recruited as neurologically healthy individuals, but were nonetheless free from ALS pathology. The ratio of male:female samples was different between cases and controls, although no clear pattern of sex bias was observed, and the longest repeats were observed almost exclusively in female controls.

The gnomAD variant catalogue is a useful tool to assess the maximum credible allele frequency of a variant^7^. However, as CNV and structural variants are not as well documented as single nucleotide variants, it is still possible to find associations between CNVs and a given phenotype that do not replicate. Samples in public databases such as gnomAD or The 1000 Genomes Project^8^ may carry large repeat alleles of risk variants^9^, but without prior evidence to strongly support variant pathogenicity, an individual might also coincidentally carry a large repeat allele. Importantly, these known limitations do not hinder our study in evaluating any proposed association of the *STMN2* CA repeat size and ALS risk.

As TDP-43 aggregation and dysfunction are central to ALS pathogenesis, lowered expression of *STMN2* could be used as a biomarker for ALS. Therefore, a variant associated both to the risk for ALS and the level of *STMN2* expression would be clinically relevant and useful. However, for a variant to be actionable, it must be strongly replicated in independent cohorts and exceed the rigorous statistical thresholds applied. While clinical genetics can and does aid in ALS management, the current results do not support the *STMN2* CA repeat as a relevant genetic risk factor.

## Data Availability

All data produced in the present study are available upon reasonable request to the authors. Please contact us if you want to use our data. We have a system in place to allow other investigators to use our data.

https://cbigr-open.loris.ca/

## Acknowledgements

JPR has received a Canadian Institutes of Health Research (CIHR) Frederick Doctoral Scholarship (FRN 159279). FA has received funding from the Fonds de Recherche du Québec– Santé (FRQS). CL has received a CIHR Vanier Graduate Scholarship (FRN 169885). PAD has received project funding from the Radala Foundation for ALS Research and jointly from the ALS Society of Canada and the Brain Canada Foundation. GAR has received funding from the ALS Society of Canada and holds a Canada Research Chair in Genetics of the Nervous System and the Wilder Penfield Chair in Neurosciences.

## Data Availability

Raw data for genome sequencing used in the study is available through Project MinE (https://www.projectmine.com/) or available upon request. Raw data for Expansionhunter variant calling is also available upon request.

## Figure Legends

**Figure S1.**
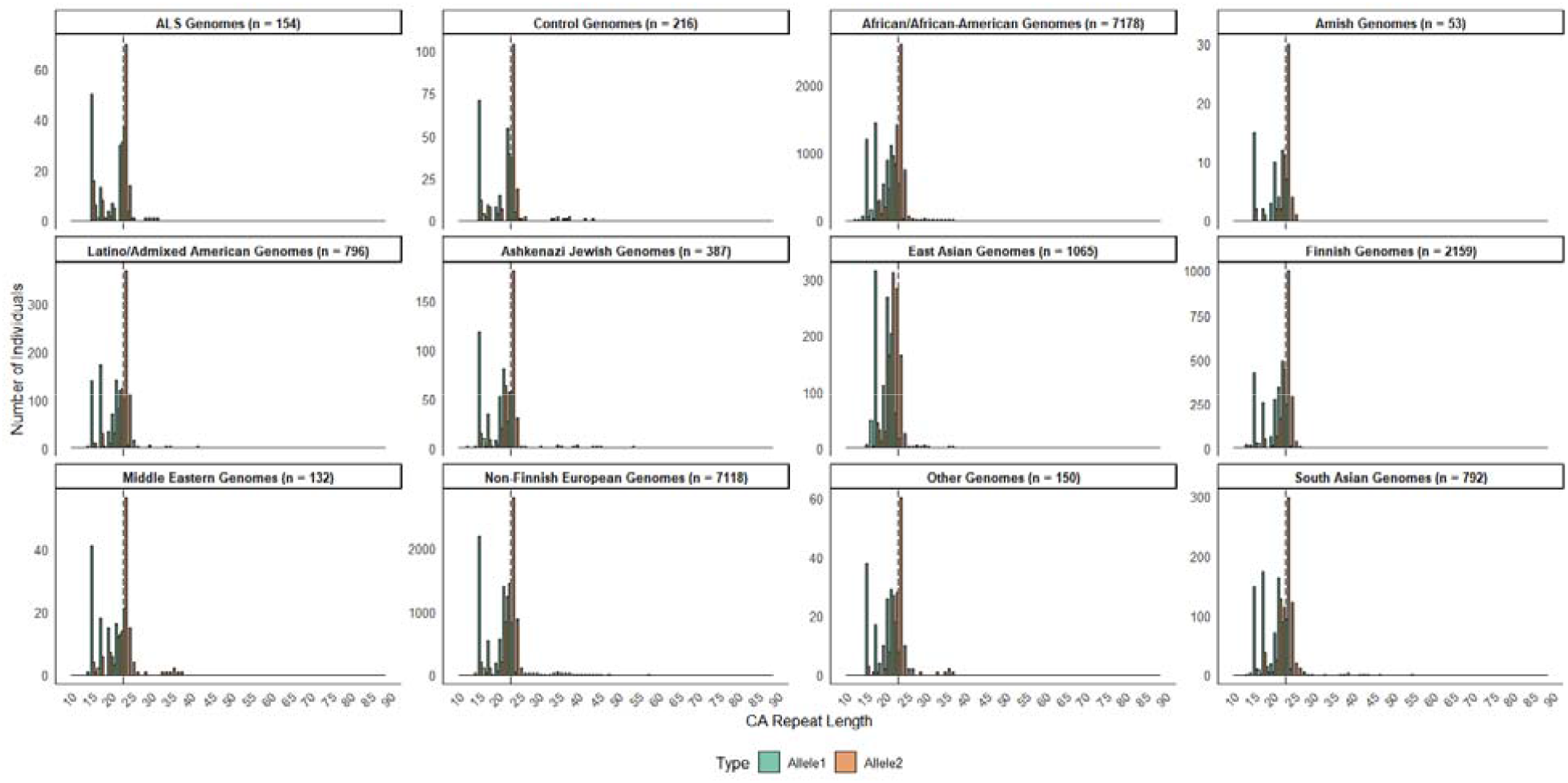
Distributions of *STMN2* CA repeat lengths for ALS, control, and individual populations from gnomAD genomes. Methods as in Figure 1, with previously reported “pathogenic” threshold of 24 repeats shown with dashed line.

**Table S1.**
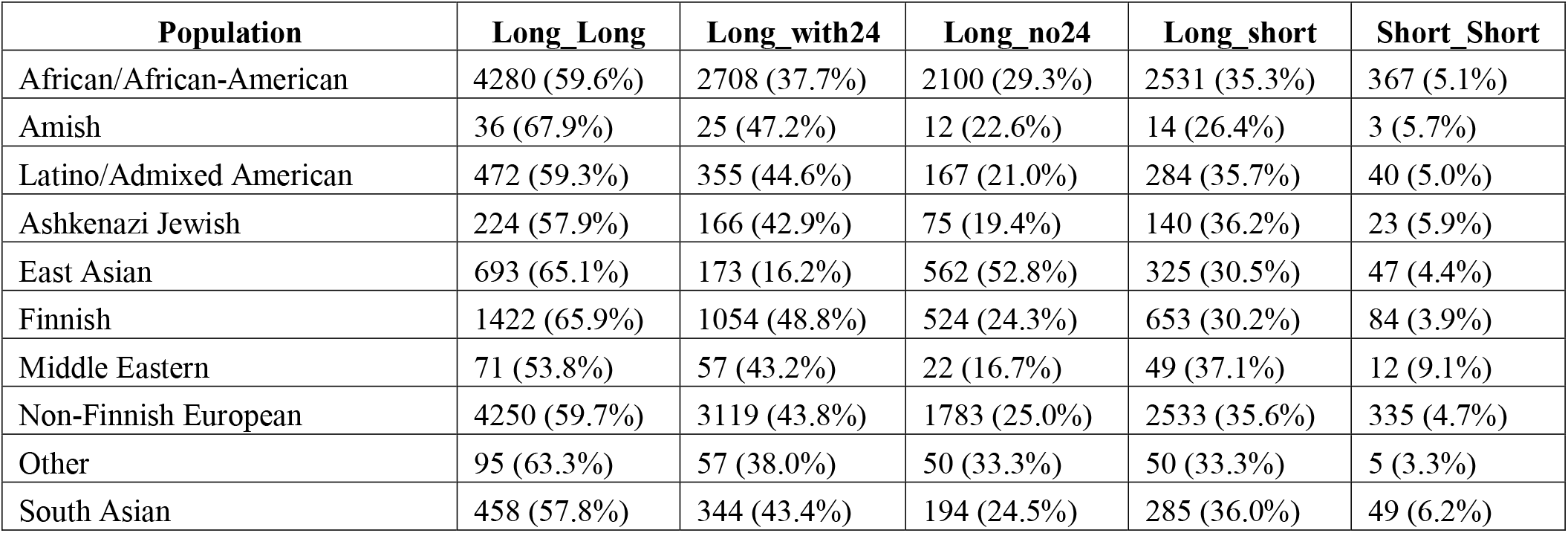
Counts and frequencies of allele combinations for each population of gnomAD genomes.

| **Population** | **Long_Long** | **Long_with24** | **Long_no24** | **Long_short** | **Short_Short** |
| --- | --- | --- | --- | --- | --- |
| African/African-American | 4280 (59.6%) | 2708 (37.7%) | 2100 (29.3%) | 2531 (35.3%) | 367 (5.1%) |
| Amish | 36 (67.9%) | 25 (47.2%) | 12 (22.6%) | 14 (26.4%) | 3 (5.7%) |
| Latino/Admixed American | 472 (59.3%) | 355 (44.6%) | 167 (21.0%) | 284 (35.7%) | 40 (5.0%) |
| Ashkenazi Jewish | 224 (57.9%) | 166 (42.9%) | 75 (19.4%) | 140 (36.2%) | 23 (5.9%) |
| East Asian | 693 (65.1%) | 173 (16.2%) | 562 (52.8%) | 325 (30.5%) | 47 (4.4%) |
| Finnish | 1422 (65.9%) | 1054 (48.8%) | 524 (24.3%) | 653 (30.2%) | 84 (3.9%) |
| Middle Eastern | 71 (53.8%) | 57 (43.2%) | 22 (16.7%) | 49 (37.1%) | 12 (9.1%) |
| Non-Finnish European | 4250 (59.7%) | 3119 (43.8%) | 1783 (25.0%) | 2533 (35.6%) | 335 (4.7%) |
| Other | 95 (63.3%) | 57 (38.0%) | 50 (33.3%) | 50 (33.3%) | 5 (3.3%) |
| South Asian | 458 (57.8%) | 344 (43.4%) | 194 (24.5%) | 285 (36.0%) | 49 (6.2%) |

**Table S2. Allele lengths and sample information for case-control cohort.** Alleles are listed in order of size, with Allele2 being the longer allele in a given individual. Sex is coded as either male (1) or female (2). Age refers to age of symptom onset or age at which a sample was donated.

## References

1 Melamed, Z. et al. Premature polyadenylation-mediated loss of stathmin-2 is a hallmark of TDP-43-dependent neurodegeneration. Nat Neurosci 22, 180–190, doi:10.1038/s41593-018-0293-z (2019).

2 Klim, J. R. et al. ALS-implicated protein TDP-43 sustains levels of STMN2, a mediator of motor neuron growth and repair. Nat Neurosci 22, 167–179, doi:10.1038/s41593-018-0300-4 (2019).

3 Theunissen, F. et al. Novel STMN2 Variant Linked to Amyotrophic Lateral Sclerosis Risk and Clinical Phenotype. Front Aging Neurosci 13, 658226, doi:10.3389/fnagi.2021.658226 (2021).

4 Karczewski, K. J. et al. The mutational constraint spectrum quantified from variation in 141,456 humans. Nature 581, 434–443, doi:10.1038/s41586-020-2308-7 (2020).

5 Dolzhenko, E. et al. ExpansionHunter: a sequence-graph-based tool to analyze variation in short tandem repeat regions. Bioinformatics 35, 4754–4756, doi:10.1093/bioinformatics/btz431 (2019).

6 Wickham, H. et al. Welcome to the Tidyverse. Journal of Open Source Software 4, doi:10.21105/joss.01686 (2019).

7 Whiffin, N. et al. Using high-resolution variant frequencies to empower clinical genome interpretation. Genet Med 19, 1151–1158, doi:10.1038/gim.2017.26 (2017).

8 Genomes Project, C. et al. A global reference for human genetic variation. Nature 526, 68–74, doi:10.1038/nature15393 (2015).

9 Akcimen, F. et al. Expanded CAG Repeats in ATXN1, ATXN2, ATXN3, and HTT in the 1000 Genomes Project. Mov Disord 36, 514–518, doi:10.1002/mds.28341 (2021).

